# What’s in a name? Protocol for a bibliometric and content analysis of rehabilitation, reablement, reactivation, and restorative health care services

**DOI:** 10.1101/2024.07.09.24309987

**Authors:** Evan MacEachern, Miranda C. Wu, Shawna Cronin, Áine Carroll, Marco Inzitari, Gaston Perman, Janet Prvu-Bettger, Michelle L. A. Nelson

**Affiliations:** Lunenfeld-Tanenbaum Research Institute, Sinai Health System, Toronto, Canada; Rehabilitation Sciences Institute, Faculty of Medicine, University of Toronto, Toronto, Canada; Department of Physical Therapy, University of Toronto, UHN, Toronto, Canada; School of Medicine, University College Dublin, Dublin, Ireland; National Rehabilitation University Hospital, Dublin, Ireland; Parc Sanitari Pere Virgili and Vall d’Hebron Institute of Research (VHIR), Barcelona, Spain; School of Health Sciences, Universitat Oberta de Catalunya, Barcelona, Spain; Universidad Hospital Italiano de Buenos Aires, Argentina; Department of Health and Rehabilitation Sciences, Temple University, Philadelphia, Pennsylvania, United States of America; Institute of Health Policy, Management and Evaluation, Dalla Lana School of Public Health, University of Toronto, Toronto, Canada

## Abstract

**Background:** Various terms are used interchangeably to describe health care services that focus on supporting functional recovery after experiencing a health event. Previous literature has identified these terms as the 4R’s: rehabilitation, reablement, reactivation, and restorative health care services. However, there lacks a clear understanding and delineation between these concepts, making it difficult to measure the efficacy of each program type. This study protocol proposes a bibliometric and content analysis to map the current scientific literature within each 4R term. **Methods:** Using a predefined search strategy, we will identify and retrieve publications from databases Scopus and PubMed between the years 1924-2024 for each 4R concept. Two independent researchers will screen articles for eligibility. Bibliometric analyses will be conducted using RStudio software and Bibliometrix and Biblioshiny extensions. Bibliometric analyses will each include a performance analysis, citation analysis, co-citation analysis, bibliographic coupling, and co-word analysis to identify key research connections and emerging trends temporally and geographically. Bibliometric indicators of interest will include total publications, yearly output, author names, and countries, among others. In addition, we will also perform a qualitative content analysis to provide a more in-depth examination of the characteristics of each program type. **Implications:** Our line of inquiry intends to clarify the similarities and differences among the 4R terms to conceptualize each definition. Findings from this study have several implications for research, practice, and policy within the 4Rs, and can overall help to delineate these concepts and facilitate decision-making and resource allocation for these health care services. This study will reveal citation patterns, research connections, and foundation themes that can inform the suitability of practice transfer and resource allocation within and between rehabilitation fields. A methodological understanding of the 4R service types can inform decision-making on the patient, healthcare professional, and system level for each service.

## Introduction

Global advancements to social and economic prosperity have led to an increase in cumulative years of life lived, stemming from greater access to health care, availability of information, and knowledge of diseases as we age.(1,2) Rising global longevity has increased the proportion of individuals who require support and services to live comfortably while effectively managing acute and chronic health adversities.(1) As a result, many systems are reimagining the design and delivery of rehabilitation services to make them more inclusive, equitable, and sustainable.(2) Global models of care have also evolved alongside patient complexities to best attend to the physical, social, and environmental considerations of service users. However, effective sharing of insights and experience to enable personalized support for rehabilitation requires a shared understanding of concepts and principles to evaluate, compare, and translate solutions into practice.

The current evidence base uses an array of terms to describe health interventions which focus on supporting functional recovery post health event, thus muddling the generalizability of implementation to the broader scope of care recipients.(3) Many health interventions staffed by comparable multidisciplinary teams will serve patient populations with similar needs, yet, such interventions are described under various labels, and distinguish themselves as nuanced models of care.(3) This utilization of ambiguous terminology to comply with jurisdictional priorities and align funding opportunities for program development may hinder the transferability of care models. Contrarily, other programs may uniformly label their health interventions, despite delivering care to diverse patient populations while implementing key methodological inconsistencies. Sims-Gould and colleagues highlighted this evidence by systematically defining reablement, reactivation, rehabilitation, and restorative health care services as the “4R’s”.(3) Often used interchangeably, the 4R’s represent time-limited, interdisciplinary, intensive programs designed to assist home-care recipients mitigate their risk of adverse health outcomes (i.e., chronic disease, functional impairment) while maximizing independence.(3) However, this primary 4R’s investigation focussed only on home-based or residence care programs.(3) While home-based care is an essential part of the rehabilitation process, there remains an opportunity to explore and conceptually understand the 4R concepts across the full pathway of rehabilitation (i.e., hospitalization, acute care, discharge, and chronic care). Other recent work has engaged in Delphi consensus methods to assemble input from global experts in the field of 4R and 4R- adjacent interventions, resulting in specific definitions now recognized as unique and independent.(4,5) Notably, a shared understanding for intermediate care (4) and an internationally accepted definition of reablement were both achieved through Delphi methods.(5)

While progress has improved the clarity of rehabilitation concepts, we must still acknowledge the challenges associated with operationalizing conceptual definitions across different health care jurisdictions.(4,5) Governmental programs and policymakers may allocate resources and funding for programs that fall under a specific call or title; the language used may especially be of interest as it may be contingent upon a program receiving the intended funding. With calls for healthy aging strategies and various governments implementing funding policies relating to home care, it is critical to examine the development of the 4R’s temporally and geographically. For example, a restorative care program will receive funding in Canada, but it must be called a reablement program to receive funding in the United Kingdom. Without an understanding of the delineation between the 4R concepts, it is difficult to measure the functional outcomes and efficacy of each program type. We posit that reablement, reactivation, rehabilitation, and restorative care services may be characterized by literature based on similar ideologies. Yet, by definition, such programs use different intensities and combinations of interdisciplinary providers (i.e., nurses, physiotherapists, occupational therapists, etc.) along the continuum of care (i.e., acute, outpatient, community reintegration) and in different settings of practice (i.e., hospitals, health care clinics, community programs). Several healthcare interventions are implemented within global health systems; however, their conceptual and operational definitions are poorly generalized, leading to limited spread and advancement of the work informing respective care models. Additionally, how these health interventions align to the concept of “rehabilitation” is not fully understood, meaning the breadth of terms used to describe reablement, reactivation, rehabilitation, and restorative interventions, and the connection between them, warrant further investigation. We seek to address a pragmatic question: Are healthcare authorities, clinicians, and rehabilitation researchers talking about the same thing?

To analyze and map the current scientific literature surrounding the 4R’s, we propose a bibliometric analysis to identify trends within and between each concept. The objectives of this bibliometric analysis are outlined to: 1) determine the accepted understanding of each model through presented definition; and 2) identify and understand different models of rehabilitative care as they relate to each other.

## Materials and Methods

We will conduct a bibliometric analysis following the methodological guidelines and employing various analyses techniques (S1 Fig.) provided by Donthu and colleagues.(6) Quantitative in nature, a bibliometric analysis examines the structural relationships between different research constituents, such as authors, countries, institutions, and topics.(6) Bibliometric analyses will focus on analyzing patterns, relationships, and trends within bibliographic data to summarize the intellectual structure of a particular field.(6) The breadth and volume of 4R’s literature suggests a bibliographic analysis is appropriate to map the current evidence base and identify foundational themes within each 4R term, as well as examine the nuances among them. To identify the structural relationships, key topics, and foundational themes for the 4R terms, we will bibliometrically analyze each domain of the 4R’s (i.e., rehabilitation, reablement, recovery, and restorative care) and provide overall comparisons between and within selected domains to translate our findings among the included terms. As summarized by Donthu and colleagues, while sharing similarities, a bibliometric analysis differs from a systematic review in its goals, scope, datasets, and analysis.(6) A systematic review aims to synthesize all relevant research evidence on a specific topic/question, while a bibliometric analysis focuses on analyzing patterns, relationships, and trends within bibliographic data, such as publications, countries, key words, authors, or journals. Our described bibliometric analysis method may appear similar to that of a systematic review as it will comprehensively examine the literature within each 4R term, however, the data extracted will be specific to the bibliometric analysis methodology outlined in Donthu and colleagues.(6) To strengthen our exploration of the 4R’s literature, we will supplement our quantitative bibliometric analysis with a qualitative content analysis. Selecting the most cited publications within each 4R field, we will perform a deductive content analysis to identify definitions used in each field and supporting details surrounding the populations and health care professionals within each 4R field. By performing a content analysis of the most influential papers in each field, we will reveal the specifics of various programs and gain a richer understanding of how these programs may relate or differ from each other. We aim to begin this bibliometric analysis and content analysis in June 2024 and anticipate completion within 12 months (i.e., June 2025).

### Participant and public involvement

Participants or the public were not involved in the design of this bibliometric analysis protocol.

#### Stage 1: Addressing the research question (RQ)

The purpose of this bibliometric analysis is to synthesize the structural characteristics of rehabilitation and rehabilitation adjacent terms as they pertain to acute and chronic disease management. Using the methodological approach of a bibliometric analysis, we seek to address the following RQs:

RQ1) How are 4R and 4R adjacent terms defined in the literature, by whom?
RQ2) What are the defining characteristics to the setting of care (e.g., primary care, acute care, inpatient rehabilitation, outpatient settings, and community-based services ?
RQ3) What are the practical applications of the identified term (e.g., case, care, problem, context, etc.), and how does this relate to the broader concept of rehabilitation?

#### Stage 2: Identifying relevant literature

To identify relevant peer-reviewed literature, our research team will create a search strategy with the assistance of an experienced librarian from the Lunenfeld-Tanenbaum Research Institute at Sinai Health System, Toronto, Ontario. We will systematically search for relevant literature in both Scopus and PubMed, as these databases are compatible with the bibliometric software we are using (described below). Our search strategy will include subject headings and text words related to the concepts of interest, such as reablement, reactivation, rehabilitation, restorative care, and any relevant subdomains using Boolean operators (AND, OR). Due to resource availability, our search will be restricted to studies available in the English language. Included articles must relate to the field of rehabilitation and rehabilitation adjacent terminology, be acquired from a peer-reviewed academic source, and be published between the years 1924-2024. We will exclude articles deemed not relevant to the field of interest that may use similar terminology (i.e. restorative care in dentistry), articles drawn from grey literature, magazines, abstract submissions, and poster presentations due to generally limited information available in these article types. A preliminary example of the search strategy is provided in S1 Appendix.

#### Stage 3: Study selection

Two reviewers, E.M. and M.W., will perform a training exercise to ensure reviewer consistency (i.e., inter-rater agreement) using a random set of 50 titles and abstracts, selecting for articles that meet our inclusion criteria. The reviewers will discuss results of the training exercise to ensure congruity of article selection and revise appropriately, ensuring the inclusion criteria is clear. The final search results will be exported to Covidence (7) to remove duplicates and screen for study inclusion. Reviewers E.M. and M.W. will independently evaluate the titles and abstracts in accordance with our review’s eligibility criteria, categorizing articles into “yes”, “no”, and “maybe” distinctions. The reviewers will examine full texts of the “yes” or “maybe” studies. This process will be repeated for each queried term. Any disagreements or conflict will be resolved through team discussions with project lead M.L.A.N.

#### Stage 4: Data extraction, analysis, & interpretation

We will broadly extract data relevant to our bibliometric analyses to assess publication trends within each field of rehabilitation and rehabilitation adjacent care. To do so, we have created pilot data extraction forms to address the proposed RQs (S1, S2, S3, S4, S5, S6 Tables). The following information for analysis will be extracted: title, abstract, full text, author name(s), country, citations, journal, DOI, references, author keywords, and index keywords (Appendix A). Additional key metrics include yearly output, publication counts, countries, citation and co-citation rates, and keyword co-occurrences. The extracted data will be analyzed using Bibliometrix software and Biblioshiny package extension(8) in R analysis software.(9) Bibliometric data will be visualized using Biblioshiny to create maps of key metrics described below. Prior to initiating our data extraction, reviewers E.M. and M.W. will test the pilot data extraction forms on five (5) articles, and if necessary, will revise the forms to ensure sufficient information is captured.

##### 4.1. Performance analysis

A performance analysis is typically used to examine the contributions that researchers provide to a given field.(10,11) For this study, the particular research constituents of interest are total publications, yearly output, author names, and countries. This will yield when and where the 4R’s have developed and how the given term used is location specific (e.g., reablement services more popular in the UK, and restorative care services more popular in Australia and New Zealand). A performance analysis will help describe timelines, locations, and contributions within each field, permitting comparison among included terms. Biblioshiny will be used to summarize the following bibliometric data: Main Information (total publications), Annual Scientific Production and Average Citations per Year (yearly output), Most Relevant Authors and Authors’ Production Over Time (author names), Countries’ Scientific Production and Countries’ Production over Time (countries). As well, the social structure between countries can be viewed using the Countries’ Collaboration World Map function.

##### 4.2. Science mapping

Science mapping is used to examine the relationships between research constituents.(10,11) We will employ the following science mapping techniques for this study: (1) citation analysis, (2) co-citation analysis, (3) bibliographic analysis, and (4) co-word analysis.

1. *Citation analysis*: assumes that citations reflect intellectual linkages between publications, and in this analysis, the impact of a publication is determined by the number of citations that it receives.(12) By employing this technique, we will be able to examine and gain an understanding of the most influential publications in each field of the included terms.(13). The data extraction form will pull the research constituents of interest (S1 Table). Within Biblioshiny, this data can be viewed through the Most Global Cited Documents and Most Local Cited References.
2. *Co-citation analysis*: assumes that publications that are cited together are similar thematically.(14) Through this analysis, we will be able to discover thematic clusters and understand the development of the foundational themes in our fields of interest.^6,15^ Data extraction will be performed using the S2, S3, and S4 Tables.
3. *Bibliographic coupling*: is similar to co-citation analysis, as it assumes that publications that share common references are similar thematically.(16,17) However, since the thematic clusters are formed based on *citing publications* and not *cited publications* like in co-citation analysis, more recent and niche publications can be identified.(6) An analysis of relationships using bibliographic analysis will allow us to explore the periodical or present development of themes in our fields of interest.(18) Examples of bibliographic coupling data extraction can be seen in S2, S3, and S4 Fig.
4. 4. *Co-word analysis*: examines the actual content of the publications, and assumes that words that frequently appear together will have a thematic relationship with one another.(6) In this case, author keywords and index keywords will be extracted for this analysis, and will allow us to explore the existing or future relationships among topics in a research field. Within the context of reablement, reactivation, rehabilitation, and restorative care, this analysis will allow us to identify common or connecting topics within and between each field.(19) Data extraction will occur utilizing the S5 and S6 Tables. An example of data visualization is shown in S5 Fig.

##### 4.3 Content Analysis

As described by Elo and Kyngäs, we will perform a content analysis following three main phases: 1) preparation, 2) organization, and 3) reporting.(20)

1. *Preparation*: Our elected deductive approach will align with our objectives of determining an accepted definition per term and determining how each existing care model relates to each other. However, we may incorporate inductive reasoning or create new categories due to the emergent nature of our content analysis. During the preparation stage, our selected unit of analysis will be the papers identified through our bibliometric analysis search. Following other bibliometric and content analysis studies,(21,22) we will select the most frequently cited publications for each 4R term. Our approach will allow our content analysis to capture the commonly cited definitions and nature of programs in each 4R term.
2. *Organization*: The chosen deductive approach requires the development of a categorization matrix. The general categories within each field we are interested in are: definitions of the concept, characteristics and demographics of populations receiving the treatment, and types of healthcare professionals providing the care. Afterward, each paper will be reviewed and coded, classifying data based on the specified categories.
3. *Reporting*: During the last stage, we will triangulate findings from our bibliometric analyses along with the content analysis to delineate the differences and similarities between and within each 4R term.

## Discussion

### Research applications

First, by identifying patterns of publication output within each field, we can gain a better understanding of how research has, and continues to progress within each domain both temporally and geographically. Examining the evolution of progress through time and locations can provide insights to the influences of co-development; for example, restorative care research output in Australia increases shortly after an increase in reablement research output in the UK. Second, this bibliometric analysis will allow us to identify research gaps between and within 4R terms. Co-citation analyses and bibliographic coupling will identify thematic clusters, revealing intersection among the fields (e.g. restorative care research often cites reablement research and vice versa). Through this, we intend to map the interconnectedness and differences between and across fields to provide a clearer representation of pragmatic rehabilitation services.

Third, bibliometric indicators and a citation analysis will evaluate the impact of research outputs and identify highly influential works within each domain. Through our bibliometric analysis, we will map collaborative networks among authors, institutions, and countries to visualize the development and emergence of research terms identified. Accordingly, we will identify key research hubs in each field and compare geographic similarities and differences among the included rehabilitation terms. As well, we will determine if the same authors and/or institutions are making meaningful contributions across multiple research fields.

Fourth, this bibliometric analysis will delineate terminology and concepts within and between rehabilitation and rehabilitation adjacent terms. Performing a co-word analysis will identify if the rehabilitation fields are using similar keywords and sharing themes. As such, we will ascertain thematic similarities should similar keywords appear within multiple fields. An examination of shared keywords can allow us to learn if rehabilitation fields share the same types of interventions and key stakeholders.

Lastly, the content analysis will supplement the bibliometric analysis and provide a more in-depth examination of the populations and programs for each field. We anticipate this qualitative component will contribute to a richer understanding of the prevalent themes and nuanced distinctions both within and across respective fields, highlighting their interrelationships.

### Research implications

This proposed bibliometric and content analysis holds several implications for research, practice, and policy in the rehabilitation space. Through the delineation of terminology and concepts, we can facilitate knowledge transfer and provide recommendations to support decision-making processes by stakeholders regarding implementation, funding, transferability across populations, and healthcare professionals between and across specialities. By analyzing the citation patterns of various rehabilitation programs we can facilitate a robust knowledge transfer among programs and health systems. If any programs are revealed to have similar citation patterns and research connections, we can determine the suitability of practice transfer from one setting to another, permitting spread and scale of highly influential programs of care to geographic areas in need. In addition, our findings could inform resource allocation within and between rehabilitation fields to address specific needs and characteristics of patient profiles and healthcare providers.

Various rehabilitation and adjacent programs will cater to distinct patient populations or feature diverse multidisciplinary teams. Understanding these comparisons can guide decisions on which patient profiles are best suited to the program of care. Such underpinnings may aid in professional development and training of rehabilitation healthcare professionals, who may be qualified to work in different types of programs should the systems align. In the case that policymakers provide funding and resources for specific programs under certain labels (i.e. “reablement” programs receive funding over “restorative” programs), stakeholders may consult the findings of this investigation to inform funding and development applications for subsequent programs. We anticipate that this study will aid in the conceptualization and formalization of each care model and highlight developments for further investigation in this space.

## Data Availability

All relevant data from this study will be made available upon study completion.

## Acknowledgements

We would like to express our gratitude to Andrea Solonsky, Information Specialist – Sinai Health System, for their expertise and support in generating the search for this manuscript.

## Supporting Information

**S1 Appendix. Preliminary Search Strategy**

**S1 Table. Citation analysis data extraction table – Summary of total number of citations and average number of citations per article.**

**S2 Table. Co-citation analysis data extraction table – Top 10 publishing journals contributing to the area of rehabilitation and rehabilitation adjacent literature.**

**S3 Table. Co-citation analysis data extraction table – Contribution of organizations based on their geographical regions.**

**S4 Table. Co-citation analysis data extraction table – Top 20 contributing organizations.**

**S5 Table. Co-word analysis data extraction table– Terms that define each cluster within the rehabilitation domain.**

**S6 Table. Co-word analysis data extraction table – Top 20 keyword occurrences in the rehabilitation term by date range.**

**S1 Fig. Figure 2 from Donthu et al. (2021) – Bibliometric analysis methods.**

**S2 Fig. Bibliographic coupling data extraction examples.**

**S3 Fig. Bibliographic coupling data extraction examples.**

**S4 Fig. Bibliographic coupling data extraction examples.**

**S5 Fig. Co-word analysis visualization – Example of keyword network.**

---

**Supplemental Figure 1.**
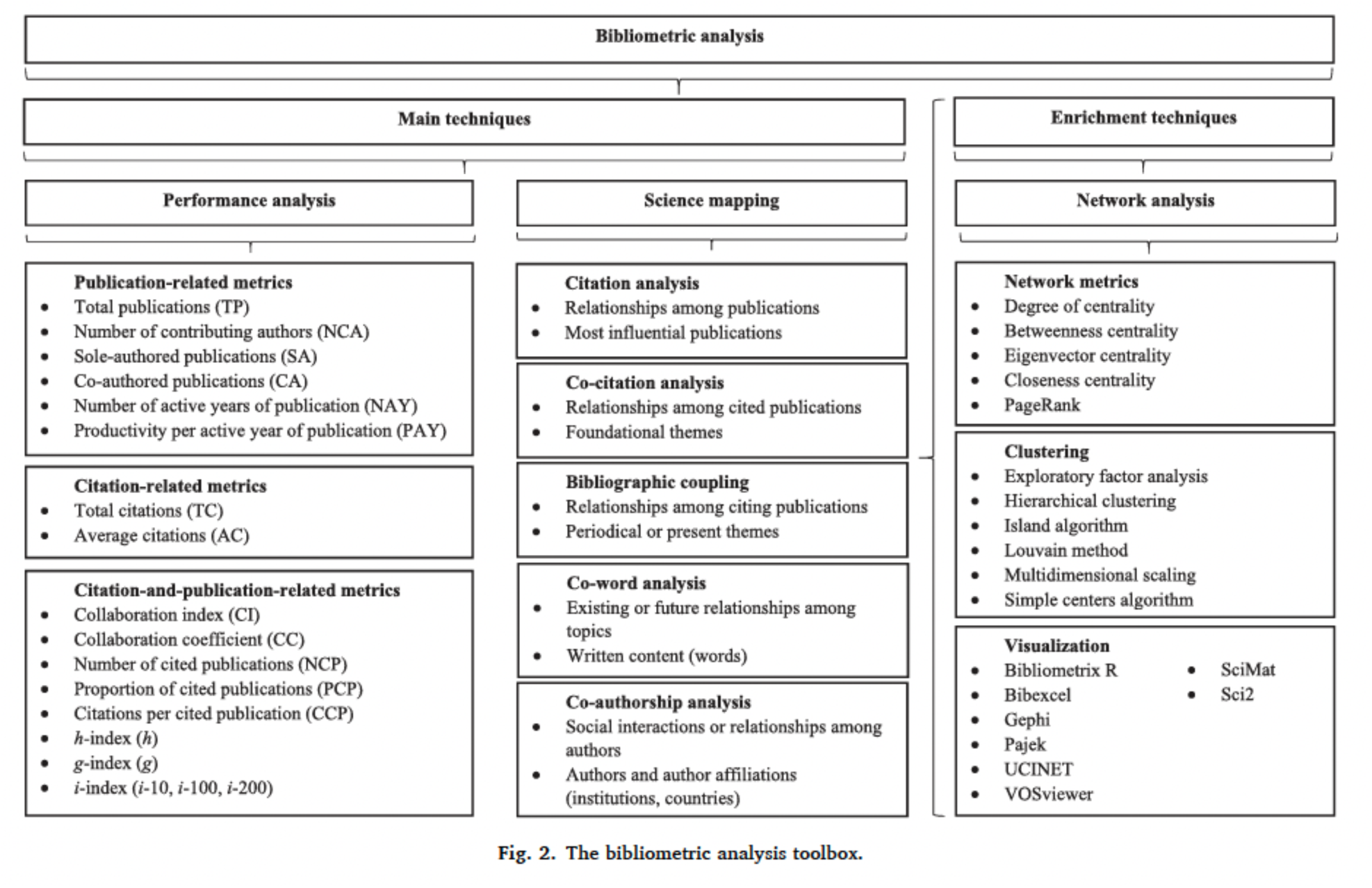
Figure 2 from Donthu et al. (2021) –Bibliometric analysis methods.

**Supplemental Figure 2.**
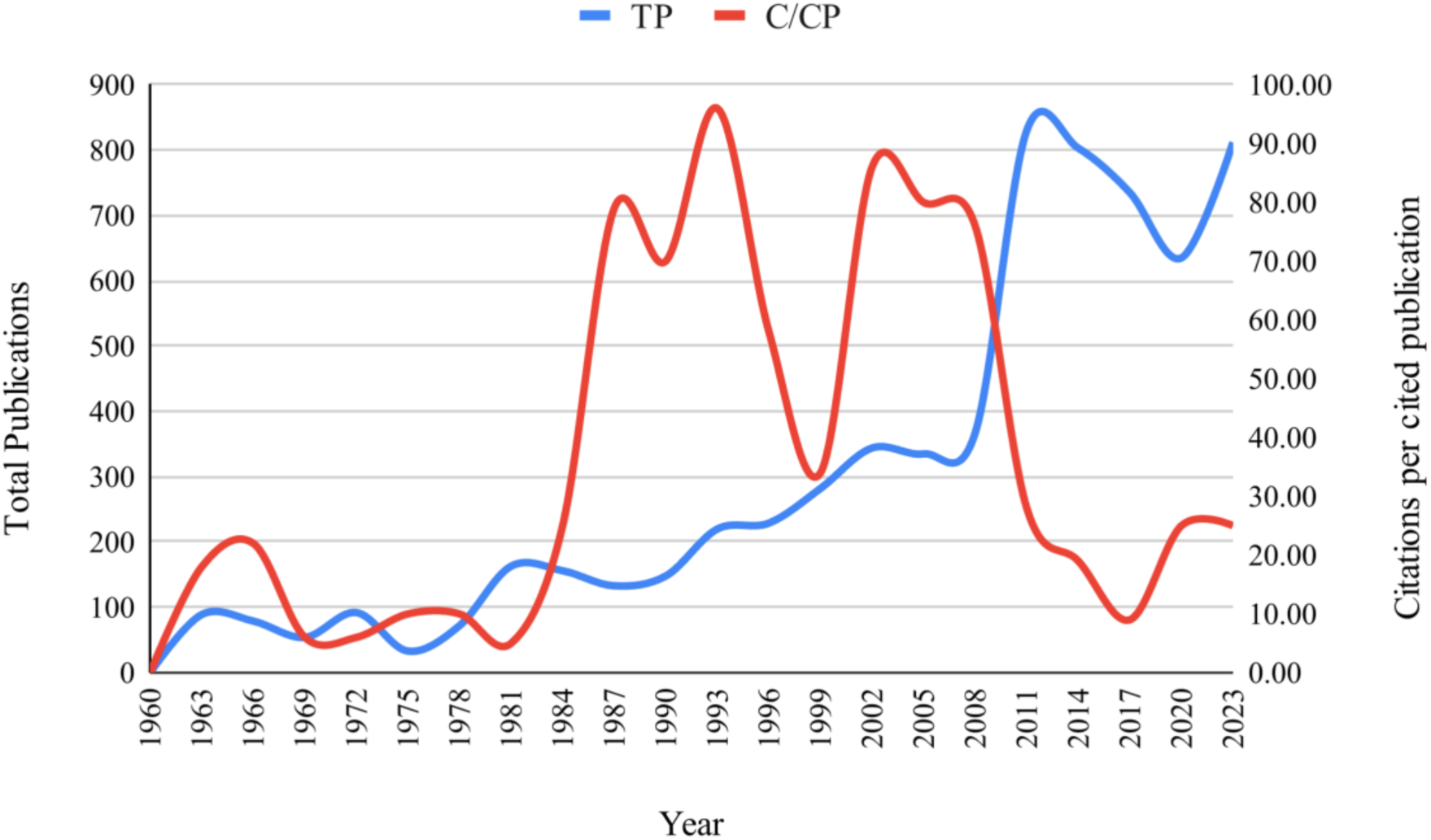
B*i*bliographic *coupling* data extraction example of publication and citation trends. TP = Total publications, C/CP = Citations per cited publication.

**Supplemental Figure 3.**
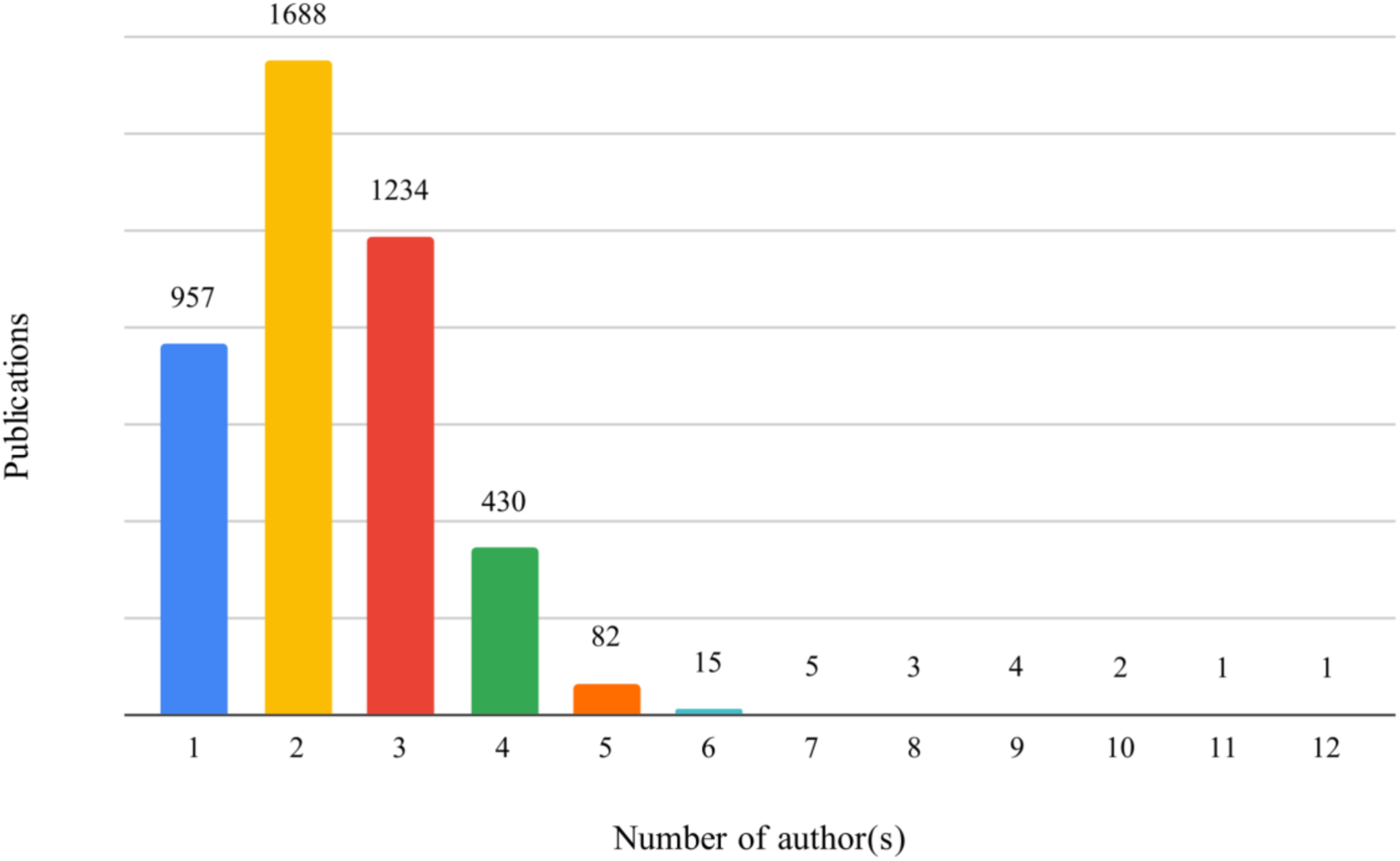
B*i*bliographic *coupling* data extraction example of distribution of publications based on number of contributing authors.

**Supplemental Figure 4.**
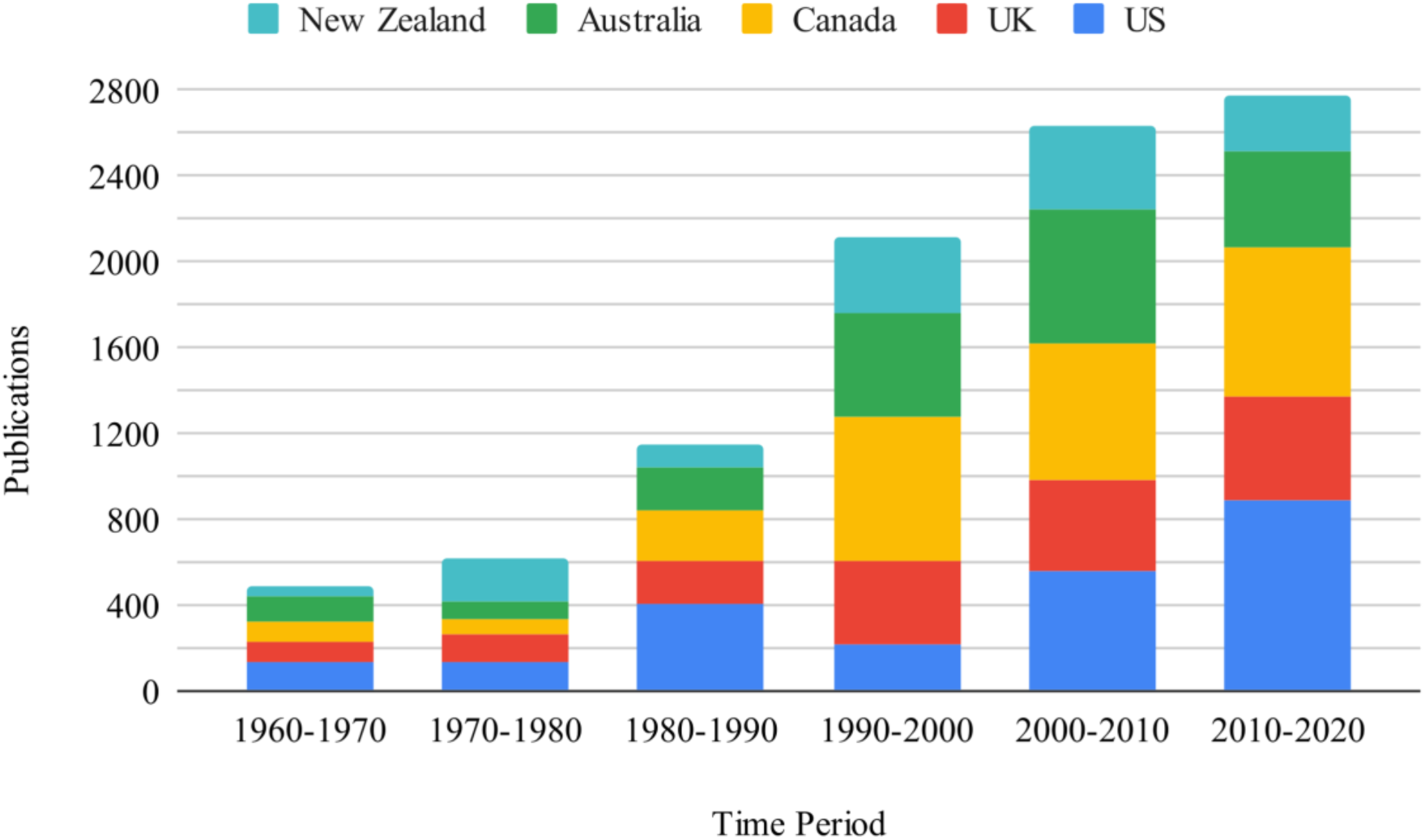
B*i*bliographic *coupling* data extraction example of the distribution of publications among five countries over time.

**Supplemental Figure 5.**
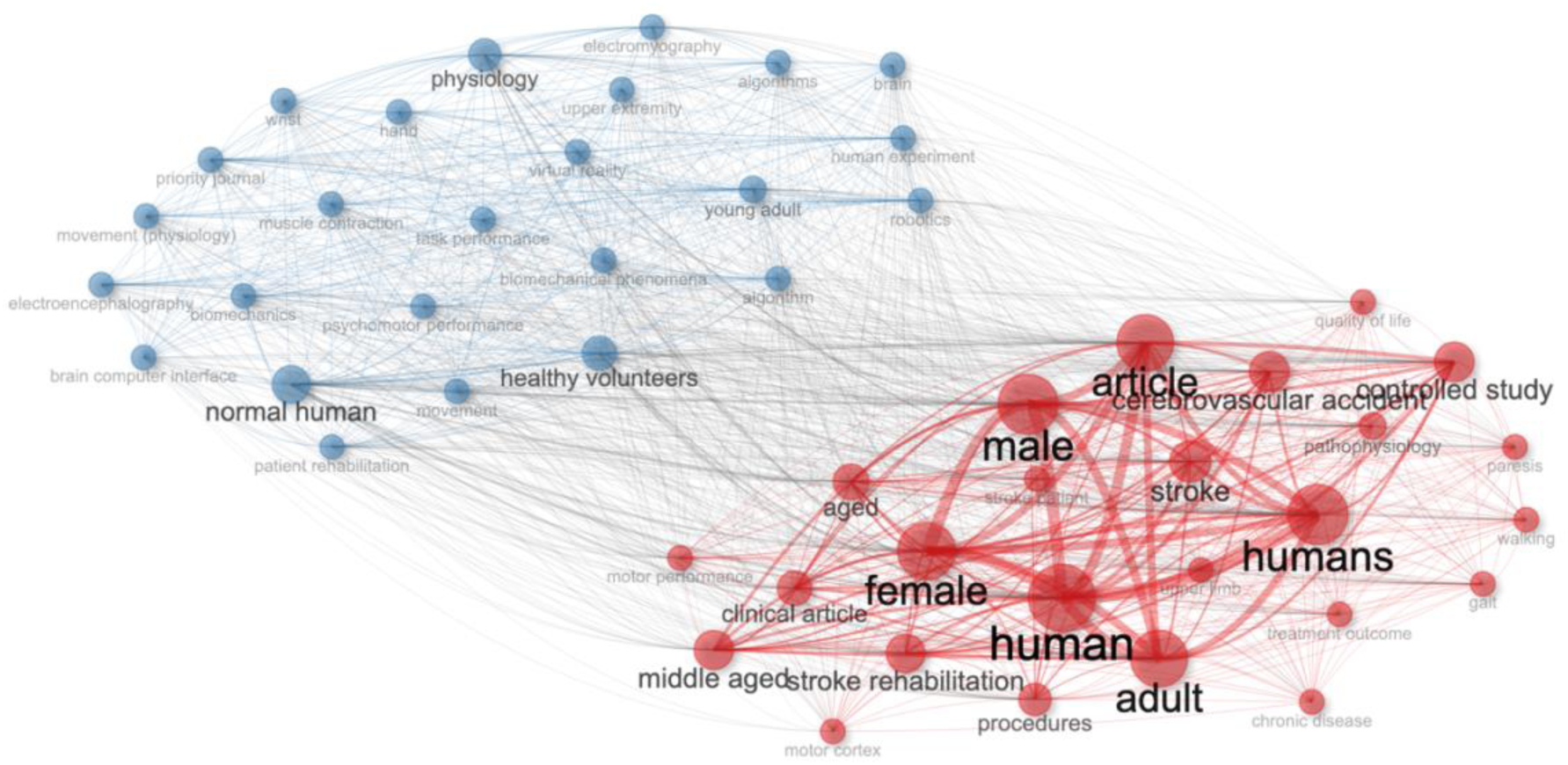
C*o-word analysis* visualization - Example of keyword network.

**Supplemental Table 1.** *Citation analysis* data extraction table –Summary of total number of citations and average number of citations per article.

| Journal | Cumulative Time Period (XX-2024) |  |  |  |  |  |
| --- | --- | --- | --- | --- | --- | --- |
|  | Total Citations |  |  | Citations/Article |  |  |
|  | # Cites | Z-Score | Rank | Average | Z-Score | Rank |
| Totals |  |  |  |  |  |  |
| Means |  |  |  |  |  |  |
| Standard deviation |  |  |  |  |  |  |
| Top 7 journals |  |  |  |  |  |  |
| Top 14 journals |  |  |  |  |  |  |
| Bottom 7 journals |  |  |  |  |  |  |

\*Additional tables could block by decade (i.e., 1970-1980, 1981-1990)

**Supplemental Table 2.**
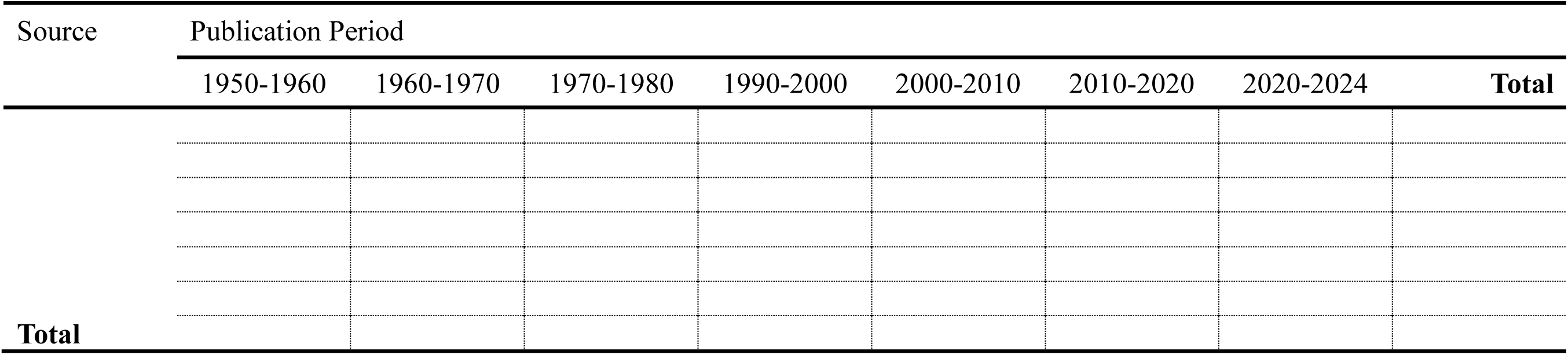
*Co-citation* analysis data extraction table –Top 10 publishing journals contributing to the area of rehabilitation and rehabilitation adjacent literature.

**Supplemental Table 3.** *Co-citation* analysis data extraction table –Contribution of organizations based on their geographical regions.

| Geographical Region | No. of papers | Percentage Contribution (%) |
| --- | --- | --- |
| Europe |  |  |
| - Northern Europe |  |  |
| - Eastern Eurasia |  |  |
| - Western Europe |  |  |
| - Southern Europe |  |  |
| North America |  |  |
| - United States |  |  |
| o Northeast US |  |  |
| o Midwest US |  |  |
| o Western US |  |  |
| o Southern US |  |  |
| - Canada |  |  |
| - Mexico |  |  |
| Asia |  |  |
| - South Asia |  |  |
| - Southeast Asia |  |  |
| - Middle East |  |  |
| Oceania |  |  |
| South America |  |  |
| Rest of the World |  |  |
| No affiliation on record |  |  |

**Supplemental Table 4.**
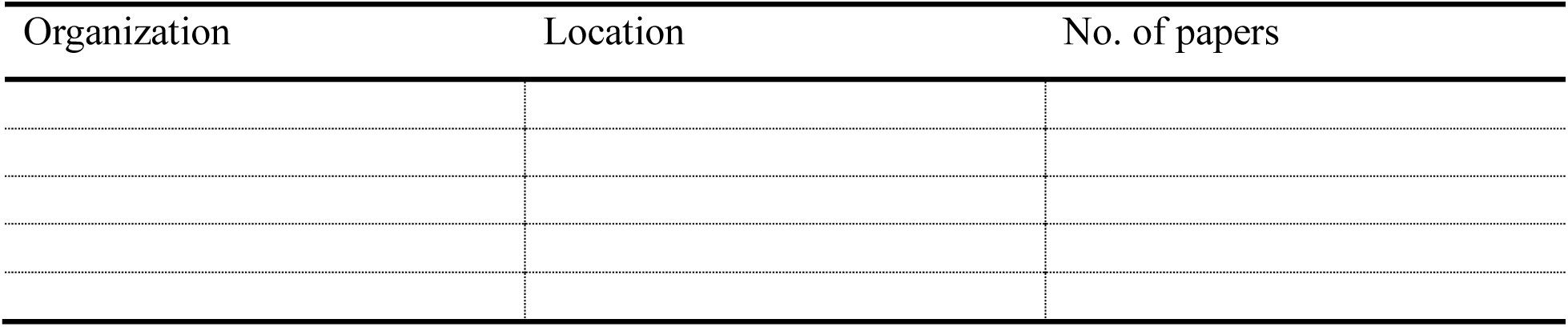
*Co-citation* analysis data extraction table –Top 20 contributing organizations.

**Supplemental Table 5.**
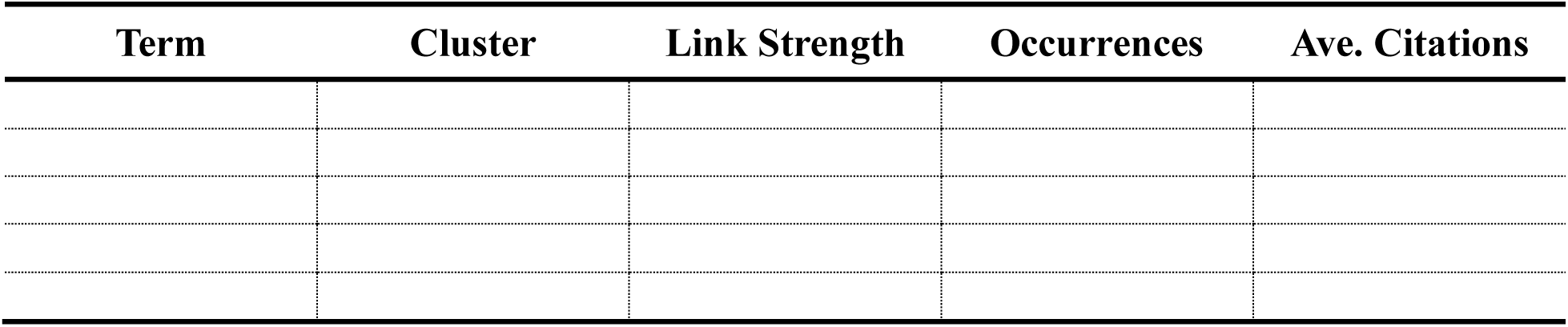
*Co-word analysis* data extraction table–Terms that define each cluster within the rehabilitation domain.

**Supplemental Table 6.**
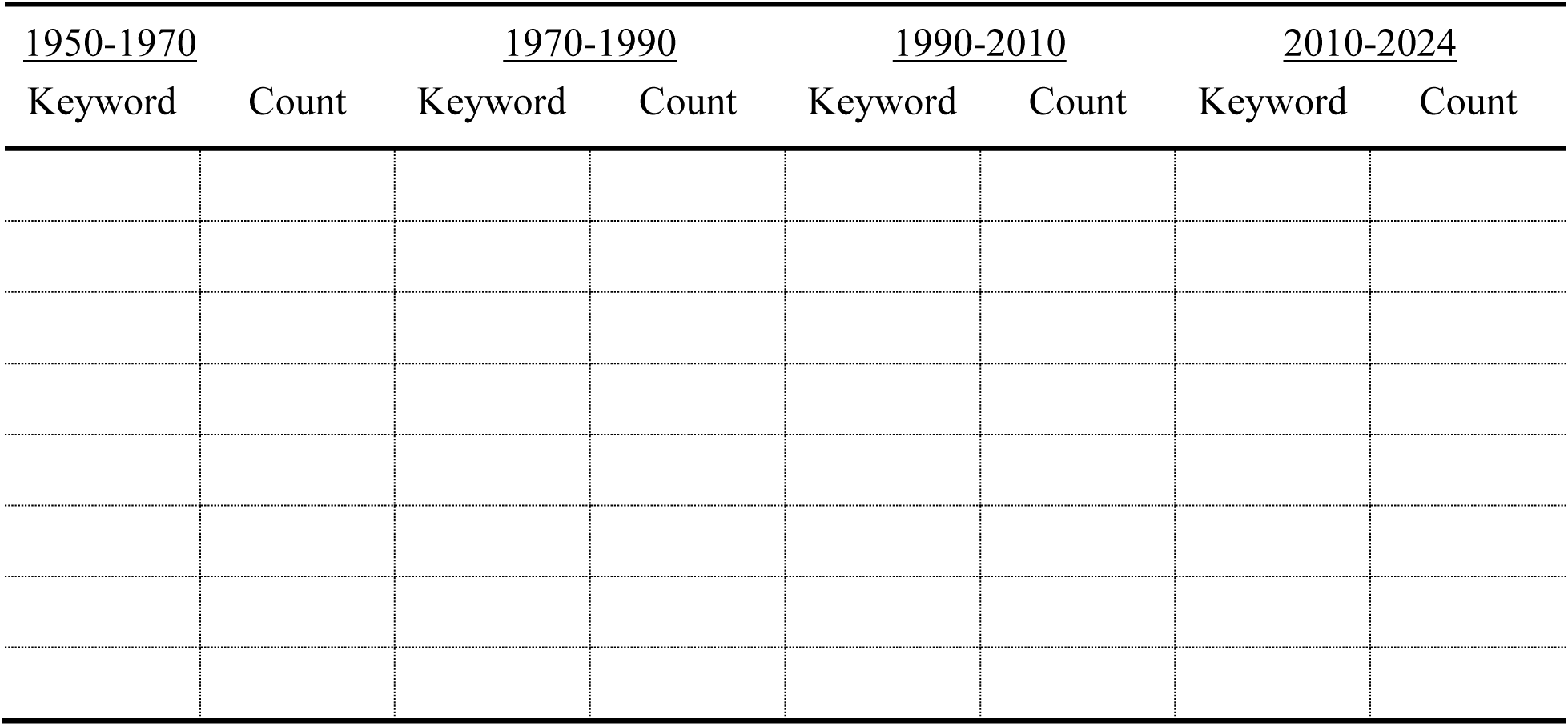
*Co-word analysis* data extraction table –Top 20 keyword occurrences in the rehabilitation term by date range.

**Supplemental Figure 1.**
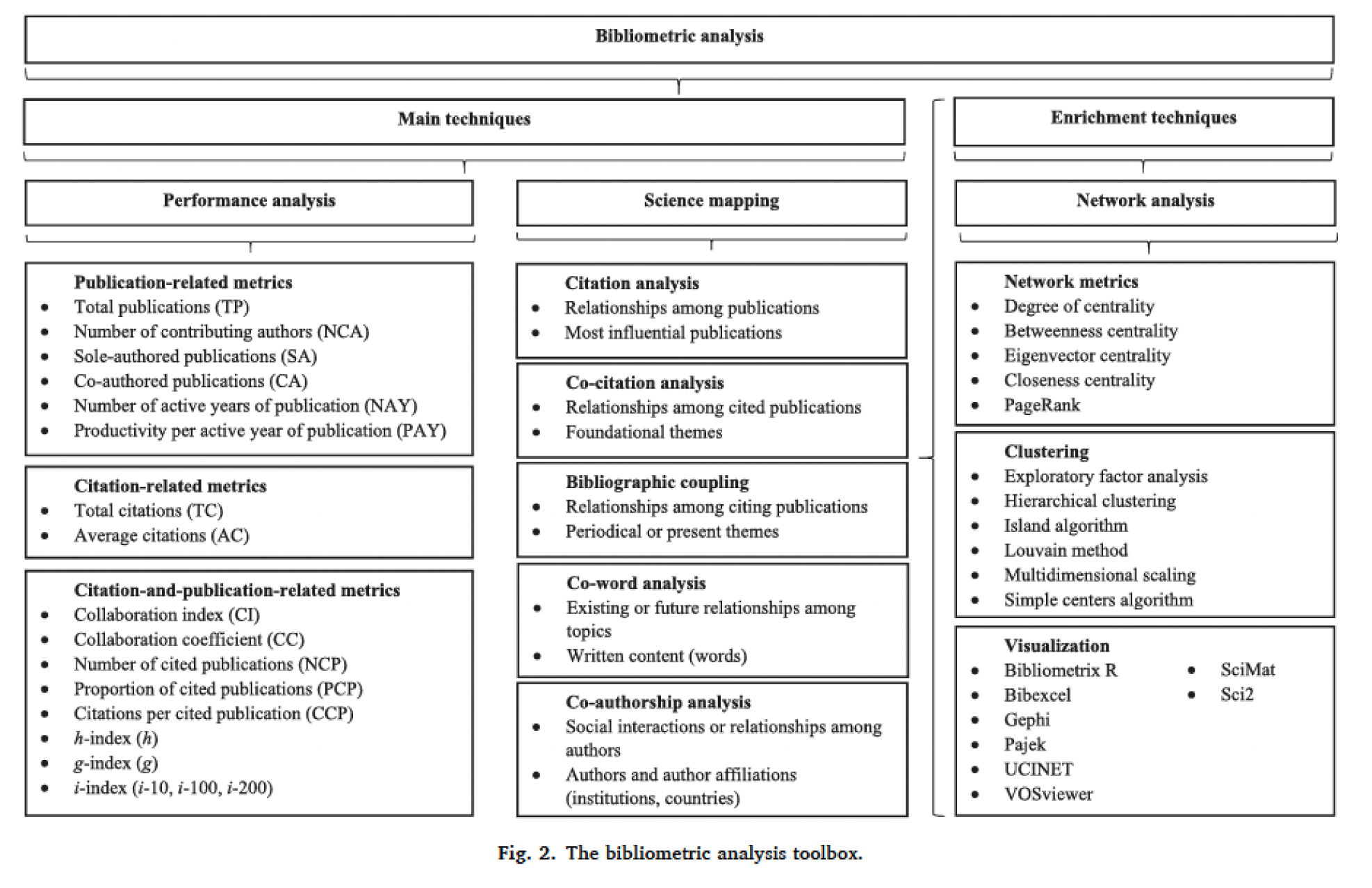
Figure 2 from Donthu et al. (2021) – How to conduct a bibliometric analysis: An overview and guidelines. Original article: https://doi.org/10.1016/j.jbusres.2021.04.070

**Supplemental Figure 2.**
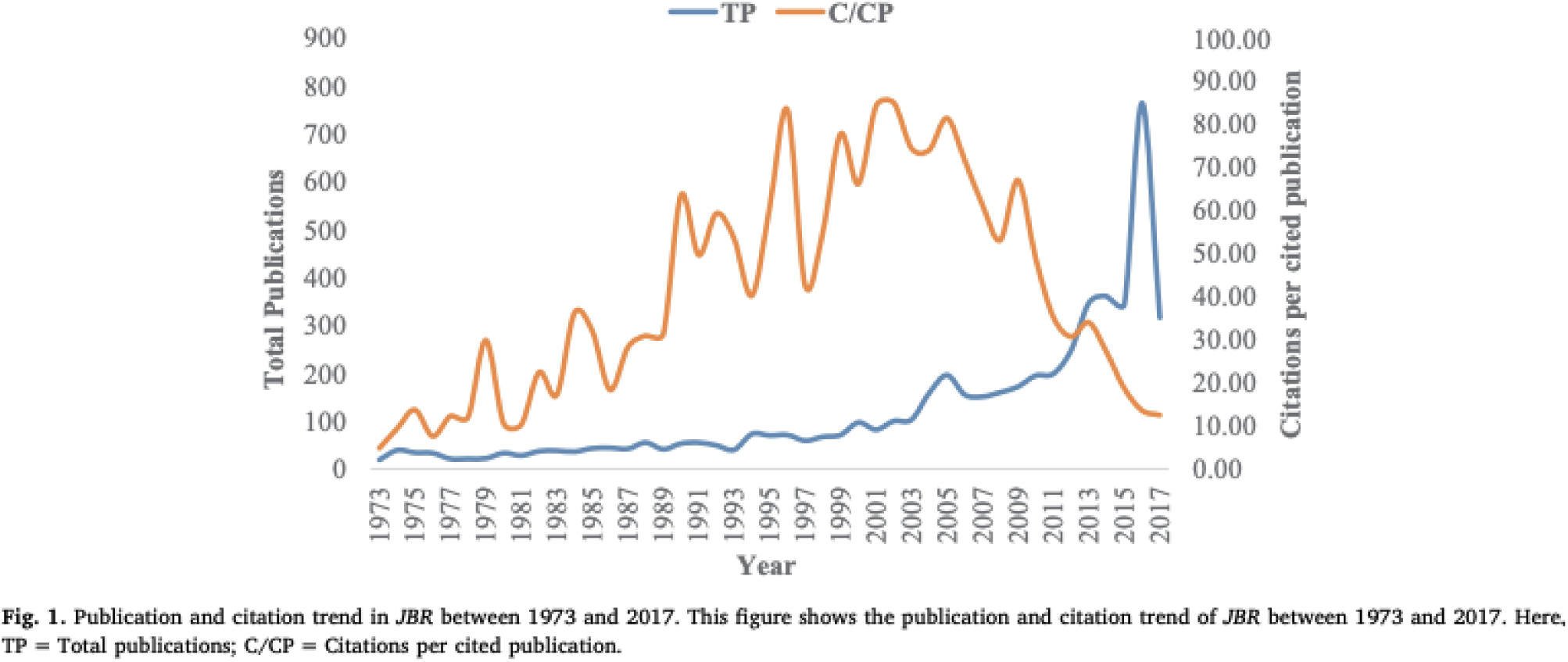
*Bibliographic coupling* data extraction examples. Original article: https://doi.org/10.1016/j.jbusres.2019.10.039

**Supplemental Figure 3.**
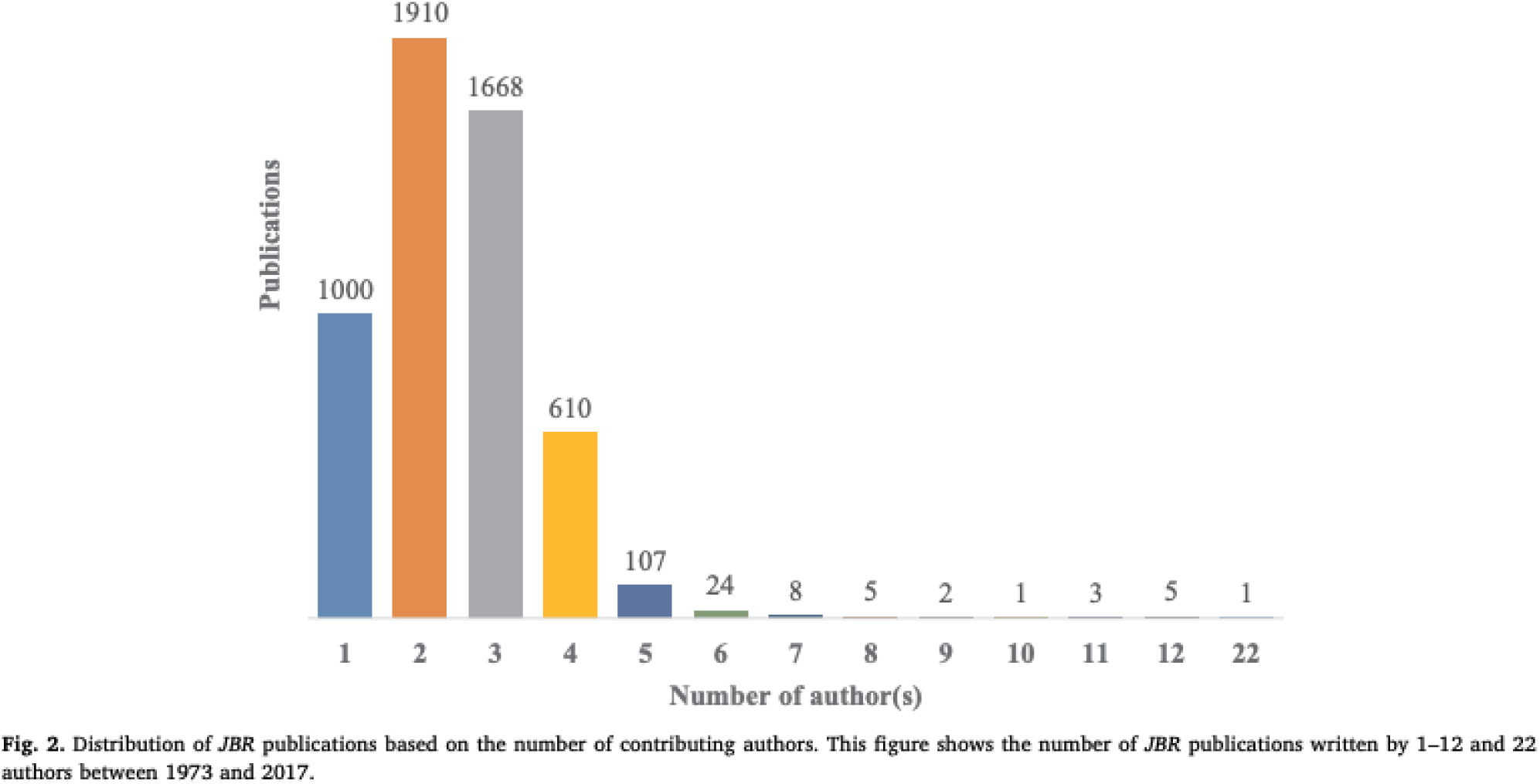
*Bibliographic coupling* data extraction examples. Original article: https://doi.org/10.1016/j.jbusres.2019.10.039

**Supplemental Figure 4.**
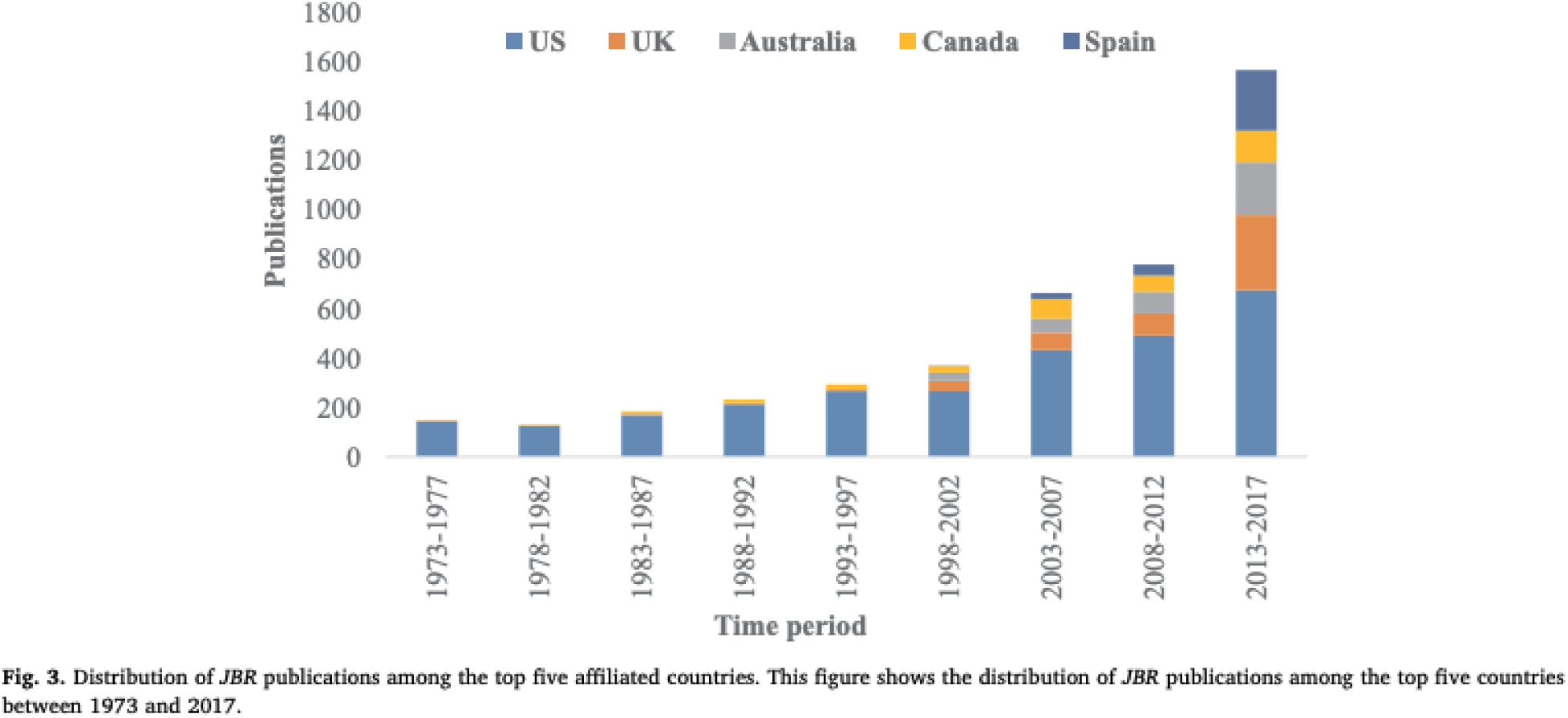
*Bibliographic coupling* data extraction examples. Original article: https://doi.org/10.1016/j.jbusres.2019.10.039

**Supplemental Figure 5.**
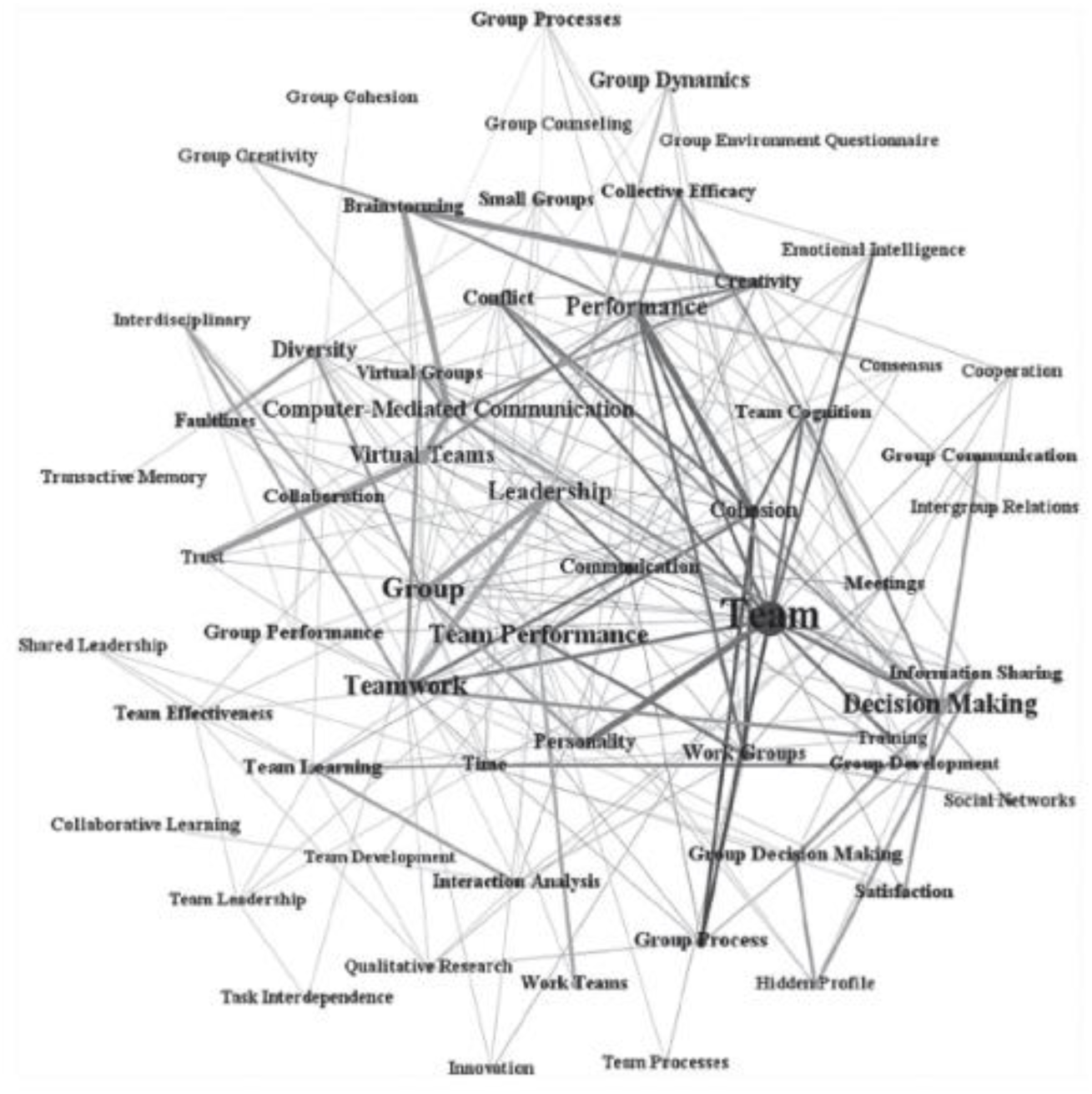
*Co-word analysis* visualization - Example of keyword network. Original article: https://doi.org/10.1177/1046496420934541

## References

1. Hay SI, Abajobir AA, Abate KH, Abbafati C, Abbas KM, Abd-Allah F, et al. Global, regional, and national disability-adjusted life-years (DALYs) for 333 diseases and injuries and healthy life expectancy (HALE) for 195 countries and territories, 1990–2016: a systematic analysis for the Global Burden of Disease Study 2016. The Lancet. 2017 Sep 16;390(10100):1260–344.

2. Stucki G, Cieza A, Melvin J. The International Classification of Functioning, Disability and Health (ICF): a unifying model for the conceptual description of the rehabilitation strategy. J Rehabil Med. 2007 May;39(4):279–85.

3. Sims-Gould J, Tong CE, Wallis-Mayer L, Ashe MC. Reablement, Reactivation, Rehabilitation and Restorative Interventions With Older Adults in Receipt of Home Care: A Systematic Review. Journal of the American Medical Directors Association. 2017 Aug;18(8):653–63.

4. Sezgin D, O’Caoimh R, O’Donovan MR, Salem MA, Kennelly S, Samaniego LL, et al. Defining the characteristics of intermediate care models including transitional care: an international Delphi study. Aging Clin Exp Res. 2020 Nov;32(11):2399–410.

5. Metzelthin SF, Rostgaard T, Parsons M, Burton E. Development of an internationally accepted definition of reablement: a Delphi study. Ageing and Society. 2022 Mar;42(3):703–18.

6. Donthu N, Kumar S, Mukherjee D, Pandey N, Lim WM. How to conduct a bibliometric analysis: An overview and guidelines. Journal of Business Research. 2021 Sep 1;133:285–96.

7. Veritas Health Innovation. Covidence systematic review software [Internet]. Melbourne, AUS; 2023 [cited 2023 Jan 16]. Available from: https://www.covidence.org/

8. Aria M, Cuccurullo C. *bibliometrix*: An R-tool for comprehensive science mapping analysis. Journal of Informetrics. 2017 Nov 1;11(4):959–75.

9. Posit Team. RStudio: Integrated Development Environment for R. [Internet]. Boston, MA: Posit Software, PBC; 2024. Available from: http://www.posit.co/

10. Cobo MJ, López-Herrera AG, Herrera-Viedma E, Herrera F. An approach for detecting, quantifying, and visualizing the evolution of a research field: A practical application to the Fuzzy Sets Theory field. Journal of Informetrics. 2011 Jan 1;5(1):146–66.

11. Ramos-Rodríguez AR, Ruíz-Navarro J. Changes in the intellectual structure of strategic management research: a bibliometric study of the Strategic Management Journal, 1980–2000. Strategic Management Journal. 2004;25(10):981–1004.

12. Appio FP, Cesaroni F, Minin A. Visualizing the structure and bridges of the intellectual property management and strategy literature: a document co-citation analysis. Scientometrics. 2014;101(1):623–61.

13. Podsakoff PM, MacKenzie SB, Bachrach DG, Podsakoff NP. The influence of management journals in the 1980s and 1990s. Strategic Management Journal. 2005;26(5):473–88.

14. Hjørland B. Facet analysis: The logical approach to knowledge organization. Information Processing & Management. 2013 Mar 1;49(2):545–57.

15. Fahimnia B, Sarkis J, Davarzani H. Green supply chain management: A review and bibliometric analysis. International Journal of Production Economics. 2015 Apr 1;162:101–14.

16. Kessler MM. Bibliographic coupling between scientific papers. American Documentation. 1963;14(1):10–25.

17. Weinberg BH. Bibliographic coupling: A review. Information Storage and Retrieval. 1974 May 1;10(5):189–96.

18. Donthu N, Kumar S, Pattnaik D. Forty-five years of *Journal of Business Research*: A bibliometric analysis. Journal of Business Research. 2020 Mar 1;109:1–14.

19. Emich KJ, Kumar S, Lu L, Norder K, Pandey N. Mapping 50 Years of Small Group Research Through Small Group Research. Small Group Research. 2020 Dec 1;51(6):659–99.

20. Elo S, Kyngäs H. The qualitative content analysis process. J Adv Nurs. 2008 Apr;62(1):107–15.

21. Salleh SZ, Bushroa AR. Bibliometric and content analysis on publications in digitization technology implementation in cultural heritage for recent five years (2016–2021). Digital Applications in Archaeology and Cultural Heritage. 2022 Jun 1;25:e00225.

22. Bhandari A. Design Thinking: from Bibliometric Analysis to Content Analysis, Current Research Trends, and Future Research Directions. Journal of the Knowledge Economy. 2022 Mar 17;14.

