## Supplementary material for "What’s in a name? Protocol for a bibliometric and content analysis of rehabilitation, reablement, reactivation, and restorative health care services": S1_Appendix

### S1 Appendix Preliminary Search Strategy

The following search strategy will be employed:

1. "crippl*"[Title/Abstract]
2. "disab*"[Title/Abstract]
3. "handicap*"[Title/Abstract]
4. "disabled persons"[MeSH Terms] OR "disabled persons"[Text Word]
5. "physical challenge"[Title/Abstract:~2]
6. "physical* challenge*"[Title/Abstract]
7. "physically challenged"[Title/Abstract]
8. "maim"[Title/Abstract] OR "maimed"[Title/Abstract] OR "maimed"[Title/Abstract] OR "maiming"[Title/Abstract]
9. "paralyze*"[Text Word] OR "paralysis"[Text Word]
10. "Paraplegia"[MeSH Terms] OR "Paraplegia"[Text Word] OR "paraplegic"[Text Word]
11. "quadriplegi*"[Text Word]
12. "traumatic brain injury"[Text Word] OR "brain injuries, traumatic"[MeSH Terms]
13. "acquired brain injury"[Text Word]
14. "physical therapy modalities"[MeSH Terms] OR "physical therapy modalities"[Text Word]
15. "physiotherap*"[Text Word]
16. "physical therapy modalities"[MeSH Terms] OR "physical therapy"[Text Word]
17. "physical therapists"[MeSH Terms] OR "physical therapist"[Text Word]
18. "occupational therapy"[MeSH Terms] OR "occupational therapy"[Text Word]
19. "ergotherap*"[Text Word]
20. "occupational therap*"[Text Word]
21. "speech and language pathologist"[Text Word]
22. "speech"[Text Word] AND "language patholog*"[Text Word]
23. "speech language patholog*"[Text Word]
24. "SLP"[Text Word]
25. "speech therapy"[MeSH Terms] OR "speech therapy"[Text Word]
26. "speech therapist"[Text Word]
27. "recreation therapy"[MeSH Terms] OR "recreation therapy"[Text Word]
28. "recreation therapist"[Text Word]
29. "therapeutic recreation"[Text Word]
30. "intermediate care facilities"[MeSH Terms] OR "intermediate care facilities"[Text Word]
31. "intermediate care"[Title/Abstract:~2]
32. "transitional care"[MeSH Terms] OR "transitional care"[Text Word] OR "transitional care"[Title/Abstract:~2]
33. "integrated care"[Title/Abstract:~2] OR "integrated care"[Text Word]
34. "Veterans"[MeSH Terms] OR "Veterans"[Text Word]
35. "re-ablement"[All Fields] AND "centre"[Text Word]
36. "reablement"[All Fields] AND "centre"[Text Word]
37. ("re-ablement"[All Fields] AND "centre"[Text Word]) OR ("reablement"[All Fields] AND "centre"[Text Word])
38. "reablement"[All Fields] AND "hospital"[Text Word]

39. "reablement"[All Fields] AND "medicine"[Text Word]
40. ("mouth"[MeSH Terms] OR "mouth"[All Fields] OR "mouths"[All Fields] OR "mouth s"[All Fields] OR "mouthed"[All Fields] OR "mouthful"[All Fields] OR "mouthfuls"[All Fields] OR "mouthing"[All Fields]) AND "reablement"[Text Word]
41. ("occupations"[MeSH Terms] OR "occupations"[All Fields] OR "vocation"[All Fields] OR "vocations"[All Fields] OR "vocational"[All Fields]) AND "reablement"[Text Word]
42. ("cardiacs"[All Fields] OR "heart"[MeSH Terms] OR "heart"[All Fields] OR "cardiac"[All Fields]) AND "reablement"[Text Word]
43. ("neurologic"[All Fields] OR "neurological"[All Fields] OR "neurologically"[All Fields]) AND "reablement"[All Fields]
44. ("psychiatrical"[All Fields] OR "psychiatrically"[All Fields] OR "psychiatrics"[All Fields] OR "psychiatry"[MeSH Terms] OR "psychiatry"[All Fields] OR "psychiatric"[All Fields]) AND "reablement"[Text Word]
45. "reablement"[All Fields] AND "nursing"[Text Word]
46. ((("physical therapy modalities"[MeSH Terms] OR "physical therapy modalities"[Text Word] OR "physiotherap*"[Text Word] OR ("physical therapy modalities"[MeSH Terms] OR "physical therapy"[Text Word]) OR ("physical therapists"[MeSH Terms] OR "physical therapist"[Text Word]) OR ("occupational therapy"[MeSH Terms] OR "occupational therapy"[Text Word]) OR "ergotherap*"[Text Word] OR "occupational therap*"[Text Word] OR "speech and language pathologist"[Text Word] OR ("Speech"[Text Word] AND "language patholog*"[Text Word]) OR ("speech therapy"[MeSH Terms] OR "speech therapy"[Text Word]) OR "speech therapist"[Text Word]) AND "Or"[All Fields]) AND ("recreation therapy"[MeSH Terms] OR "recreation therapy"[Text Word])) OR "recreation therapist"[Text Word] OR "therapeutic recreation"[Text Word] OR (("mouth"[MeSH Terms] OR "mouth"[All Fields] OR "mouths"[All Fields] OR "mouth s"[All Fields] OR "mouthed"[All Fields] OR "mouthful"[All Fields] OR "mouthfuls"[All Fields] OR "mouthing"[All Fields]) AND "reablement"[Text Word]) OR (("occupations"[MeSH Terms] OR "occupations"[All Fields] OR "vocation"[All Fields] OR "vocations"[All Fields] OR "vocational"[All Fields]) AND "reablement"[Text Word]) OR (("cardiacs"[All Fields] OR "heart"[MeSH Terms] OR "heart"[All Fields] OR "cardiac"[All Fields]) AND "reablement"[Text Word]) OR (("neurologic"[All Fields] OR "neurological"[All Fields] OR "neurologically"[All Fields]) AND "reablement"[All Fields]) OR (("psychiatrical"[All Fields] OR "psychiatrically"[All Fields] OR "psychiatrics"[All Fields] OR "psychiatry"[MeSH Terms] OR "psychiatry"[All Fields] OR "psychiatric"[All Fields]) AND "reablement"[Text Word]) OR ("reablement"[All Fields] AND "nursing"[Text Word])
47. ((("physical therapy modalities"[MeSH Terms] OR "physical therapy modalities"[Text Word] OR "physiotherap*"[Text Word] OR ("physical therapy modalities"[MeSH Terms] OR "physical therapy"[Text Word]) OR ("physical therapists"[MeSH Terms] OR "physical therapist"[Text Word]) OR ("occupational therapy"[MeSH Terms] OR "occupational therapy"[Text Word]) OR "ergotherap*"[Text Word] OR "occupational therap*"[Text Word] OR "speech and language pathologist"[Text Word] OR ("Speech"[Text Word] AND "language patholog*"[Text Word]) OR "speech language patholog*"[Text Word] OR "SLP"[Text Word] OR ("speech therapy"[MeSH Terms] OR "speech therapy"[Text Word]) OR "speech therapist"[Text Word]) AND "Or"[All Fields]) AND ("recreation therapy"[MeSH Terms] OR "recreation therapy"[Text Word])) OR

"recreation therapist"[Text Word] OR "therapeutic recreation"[Text Word] OR (("mouth"[MeSH Terms] OR "mouth"[All Fields] OR "mouths"[All Fields] OR "mouth s"[All Fields] OR "mouthed"[All Fields] OR "mouthful"[All Fields] OR "mouthfuls"[All Fields] OR "mouthing"[All Fields]) AND "reablement"[Text Word]) OR (("occupations"[MeSH Terms] OR "occupations"[All Fields] OR "vocation"[All Fields] OR "vocations"[All Fields] OR "vocational"[All Fields]) AND "reablement"[Text Word]) OR (("cardiacs"[All Fields] OR "heart"[MeSH Terms] OR "heart"[All Fields] OR "cardiac"[All Fields]) AND "reablement"[Text Word]) OR (("neurologic"[All Fields] OR "neurological"[All Fields] OR "neurologically"[All Fields]) AND "reablement"[All Fields]) OR (("psychiatrical"[All Fields] OR "psychiatrically"[All Fields] OR "psychiatrics"[All Fields] OR "psychiatry"[MeSH Terms] OR "psychiatry"[All Fields] OR "psychiatric"[All Fields]) AND "reablement"[Text Word]) OR ("reablement"[All Fields] AND "nursing"[Text Word])

48. ("reablement"[All Fields] AND "medicine"[Text Word]) OR (("mouth"[MeSH Terms] OR "mouth"[All Fields] OR "mouths"[All Fields] OR "mouth s"[All Fields] OR "mouthed"[All Fields] OR "mouthful"[All Fields] OR "mouthfuls"[All Fields] OR "mouthing"[All Fields]) AND "reablement"[Text Word]) OR (("occupations"[MeSH Terms] OR "occupations"[All Fields] OR "vocation"[All Fields] OR "vocations"[All Fields] OR "vocational"[All Fields]) AND "reablement"[Text Word]) OR (("cardiacs"[All Fields] OR "heart"[MeSH Terms] OR "heart"[All Fields] OR "cardiac"[All Fields]) AND "reablement"[Text Word]) OR (("neurologic"[All Fields] OR "neurological"[All Fields] OR "neurologically"[All Fields]) AND "reablement"[All Fields]) OR (("neurologic"[All Fields] OR "neurological"[All Fields] OR "neurologically"[All Fields]) AND "reablement"[All Fields]) OR (("psychiatrical"[All Fields] OR "psychiatrically"[All Fields] OR "psychiatrics"[All Fields] OR "psychiatry"[MeSH Terms] OR "psychiatry"[All Fields] OR "psychiatric"[All Fields]) AND "reablement"[Text Word]) OR ("reablement"[All Fields] AND "nursing"[Text Word]) OR (("stroke"[MeSH Terms] OR "stroke"[All Fields] OR "strokes"[All Fields] OR "stroke s"[All Fields]) AND "reablement"[Text Word])
49. "amputation"[Text Word] OR "amputation, surgical"[MeSH Terms] OR "Veterans"[MeSH Terms] OR "Veterans"[Text Word]
50. "amputation"[Text Word] OR "amputation, surgical"[MeSH Terms]
51. ("re-ablement"[All Fields] AND "centre"[Text Word]) OR ("reablement"[All Fields] AND "centre"[Text Word]) OR ("reablement"[All Fields] AND "hospital"[Text Word]) OR ("integrated care"[Title/Abstract:~2] OR "integrated care"[Text Word]) OR ("transitional care"[MeSH Terms] OR "transitional care"[Text Word] OR "transitional care"[Title/Abstract:~2]) OR ("intermediate care facilities"[MeSH Terms] OR "intermediate care facilities"[Text Word]) OR "reablement care"[Text Word]
52. (("re-ablement"[All Fields] AND "centre"[Text Word]) OR ("reablement"[All Fields] AND "centre"[Text Word]) OR ("reablement"[All Fields] AND "hospital"[Text Word]) OR ("integrated care"[Title/Abstract:~2] OR "integrated care"[Text Word]) OR ("transitional care"[MeSH Terms] OR "transitional care"[Text Word] OR "transitional care"[Title/Abstract:~2]) OR ("intermediate care facilities"[MeSH Terms] OR "intermediate care facilities"[Text Word]) OR "reablement care"[Text Word]) AND ("amputation"[Text Word] OR "amputation, surgical"[MeSH Terms] OR ("Veterans"[MeSH Terms] OR "Veterans"[Text Word]) OR ("crippl*"[Title/Abstract] OR

"disab*"[Title/Abstract] OR "handicap*"[Title/Abstract] OR ("disabled persons"[MeSH Terms] OR "disabled persons"[Text Word]) OR "physical challenge"[Title/Abstract:~2] OR "physical* challenge*"[Title/Abstract] OR "physically challenged"[Title/Abstract] OR ("maim"[Title/Abstract] OR "maimed"[Title/Abstract] OR "maimed"[Title/Abstract] OR "maiming"[Title/Abstract]) OR ("paralyze*"[Text Word] OR "paralysis"[Text Word]) OR ("Paraplegia"[MeSH Terms] OR "Paraplegia"[Text Word] OR "paraplegic"[Text Word]) OR "quadriplegi*"[Text Word] OR ("traumatic brain injury"[Text Word] OR "brain injuries, traumatic"[MeSH Terms]) OR "acquired brain injury"[Text Word])) AND (("reablement"[All Fields] AND "medicine"[Text Word]) OR (("mouth"[MeSH Terms] OR "mouth"[All Fields] OR "mouths"[All Fields] OR "mouth s"[All Fields] OR "mouthed"[All Fields] OR "mouthful"[All Fields] OR "mouthfuls"[All Fields] OR "mouthing"[All Fields]) AND "reablement"[Text Word]) OR (("occupations"[MeSH Terms] OR "occupations"[All Fields] OR "vocation"[All Fields] OR "vocations"[All Fields] OR "vocational"[All Fields]) AND "reablement"[Text Word]) OR (("cardiacs"[All Fields] OR "heart"[MeSH Terms] OR "heart"[All Fields] OR "cardiac"[All Fields]) AND "reablement"[Text Word]) OR (("neurologic"[All Fields] OR "neurological"[All Fields] OR "neurologically"[All Fields]) AND "reablement"[All Fields]) OR (("neurologic"[All Fields] OR "neurological"[All Fields] OR "neurologically"[All Fields]) AND "reablement"[All Fields]) OR (("psychiatrical"[All Fields] OR "psychiatrically"[All Fields] OR "psychiatrics"[All Fields] OR "psychiatry"[MeSH Terms] OR "psychiatry"[All Fields] OR "psychiatric"[All Fields]) AND "reablement"[Text Word]) OR ("reablement"[All Fields] AND "nursing"[Text Word]) OR (("stroke"[MeSH Terms] OR "stroke"[All Fields] OR "strokes"[All Fields] OR "stroke s"[All Fields]) AND "reablement"[Text Word]) OR (((("physical therapy modalities"[MeSH Terms] OR "physical therapy modalities"[Text Word] OR "physiotherap*"[Text Word] OR ("physical therapy modalities"[MeSH Terms] OR "physical therapy"[Text Word]) OR ("physical therapists"[MeSH Terms] OR "physical therapist"[Text Word]) OR ("occupational therapy"[MeSH Terms] OR "occupational therapy"[Text Word]) OR "ergotherap*"[Text Word] OR "occupational therap*"[Text Word] OR "speech and language pathologist"[Text Word] OR ("Speech"[Text Word] AND "language patholog*"[Text Word]) OR "speech language patholog*"[Text Word] OR "SLP"[Text Word] OR ("speech therapy"[MeSH Terms] OR "speech therapy"[Text Word]) OR "speech therapist"[Text Word]) AND "Or"[All Fields]) AND ("recreation therapy"[MeSH Terms] OR "recreation therapy"[Text Word])) OR "recreation therapist"[Text Word] OR "therapeutic recreation"[Text Word]))

53. "amputation"[Text Word] OR "amputation, surgical"[MeSH Terms] OR "Veterans"[MeSH Terms] OR "Veterans"[Text Word] OR "crippl*"[Title/Abstract] OR "disab*"[Title/Abstract] OR "handicap*"[Title/Abstract] OR "disabled persons"[MeSH Terms] OR "disabled persons"[Text Word] OR "physical challenge"[Title/Abstract:~2] OR "physical* challenge*"[Title/Abstract] OR "physically challenged"[Title/Abstract] OR "maim"[Title/Abstract] OR "maimed"[Title/Abstract] OR "maimed"[Title/Abstract] OR "maiming"[Title/Abstract] OR "paralyze*"[Text Word] OR "paralysis"[Text Word] OR "Paraplegia"[MeSH Terms] OR "Paraplegia"[Text Word] OR "paraplegic"[Text Word] OR "quadriplegi*"[Text Word] OR "traumatic brain injury"[Text Word] OR "brain injuries, traumatic"[MeSH Terms] OR "acquired brain injury"[Text Word]

54. ("reablement"[All Fields] AND "medicine"[Text Word]) OR (("mouth"[MeSH Terms] OR "mouth"[All Fields] OR "mouths"[All Fields] OR "mouth s"[All Fields] OR "mouthed"[All Fields] OR "mouthful"[All Fields] OR "mouthfuls"[All Fields] OR "mouthing"[All Fields]) AND "reablement"[Text Word]) OR (("occupation"[MeSH Terms] OR "occupation"[All Fields] OR "vocation"[All Fields] OR "vocations"[All Fields] OR "vocational"[All Fields]) AND "reablement"[Text Word]) OR (("cardiacs"[All Fields] OR "heart"[MeSH Terms] OR "heart"[All Fields] OR "cardiac"[All Fields]) AND "reablement"[Text Word]) OR (("neurologic"[All Fields] OR "neurological"[All Fields] OR "neurologically"[All Fields]) AND "reablement"[All Fields]) OR (("neurologic"[All Fields] OR "neurological"[All Fields] OR "neurologically"[All Fields]) AND "reablement"[All Fields]) OR (("psychiatrical"[All Fields] OR "psychiatrically"[All Fields] OR "psychiatrics"[All Fields] OR "psychiatry"[MeSH Terms] OR "psychiatry"[All Fields] OR "psychiatric"[All Fields]) AND "reablement"[Text Word]) OR ("reablement"[All Fields] AND "nursing"[Text Word]) OR (("stroke"[MeSH Terms] OR "stroke"[All Fields] OR "strokes"[All Fields] OR "stroke s"[All Fields]) AND "reablement"[Text Word]) OR (((("physical therapy modalities"[MeSH Terms] OR "physical therapy modalities"[Text Word] OR "physiotherap*"[Text Word] OR ("physical therapy modalities"[MeSH Terms] OR "physical therapy"[Text Word]) OR ("physical therapists"[MeSH Terms] OR "physical therapist"[Text Word]) OR ("occupational therapy"[MeSH Terms] OR "occupational therapy"[Text Word]) OR "ergotherap*"[Text Word] OR "occupational therap*"[Text Word] OR "speech and language pathologist"[Text Word] OR ("Speech"[Text Word] AND "language patholog*"[Text Word]) OR "speech language patholog*"[Text Word] OR "SLP"[Text Word] OR ("speech therapy"[MeSH Terms] OR "speech therapy"[Text Word]) OR "speech therapist"[Text Word]) AND "Or"[All Fields]) AND ("recreation therapy"[MeSH Terms] OR "recreation therapy"[Text Word])) OR "recreation therapist"[Text Word] OR "therapeutic recreation"[Text Word])
55. ((("physical therapy modalities"[MeSH Terms] OR "physical therapy modalities"[Text Word] OR "physiotherap*"[Text Word] OR ("physical therapy modalities"[MeSH Terms] OR "physical therapy"[Text Word]) OR ("physical therapists"[MeSH Terms] OR "physical therapist"[Text Word]) OR ("occupational therapy"[MeSH Terms] OR "occupational therapy"[Text Word]) OR "ergotherap*"[Text Word] OR "occupational therap*"[Text Word] OR "speech and language pathologist"[Text Word] OR ("Speech"[Text Word] AND "language patholog*"[Text Word]) OR "speech language patholog*"[Text Word] OR "SLP"[Text Word] OR ("speech therapy"[MeSH Terms] OR "speech therapy"[Text Word]) OR "speech therapist"[Text Word]) AND "Or"[All Fields]) AND ("recreation therapy"[MeSH Terms] OR "recreation therapy"[Text Word])) OR "recreation therapist"[Text Word] OR "therapeutic recreation"[Text Word]
56. "crippl*"[Title/Abstract] OR "disab*"[Title/Abstract] OR "handicap*"[Title/Abstract] OR "disabled persons"[MeSH Terms] OR "disabled persons"[Text Word] OR "physical challenge"[Title/Abstract:~2] OR "physical* challenge*"[Title/Abstract] OR "physically challenged"[Title/Abstract] OR "maim"[Title/Abstract] OR "maimed"[Title/Abstract] OR "maimed"[Title/Abstract] OR "maiming"[Title/Abstract] OR "paralyze*"[Text Word] OR "paralysis"[Text Word] OR "Paraplegia"[MeSH Terms] OR "Paraplegia"[Text Word] OR "paraplegic"[Text Word] OR "quadriplegi*"[Text Word] OR "traumatic brain injury"[Text

Word] OR "brain injuries, traumatic"[MeSH Terms] OR "acquired brain injury"[Text Word]

57. ("amputation"[Text Word] OR "amputation, surgical"[MeSH Terms] OR ("Veterans"[MeSH Terms] OR "Veterans"[Text Word]) OR ("crippl*"[Title/Abstract] OR "disab*"[Title/Abstract] OR "handicap*"[Title/Abstract] OR ("disabled persons"[MeSH Terms] OR "disabled persons"[Text Word]) OR "physical challenge"[Title/Abstract:~2] OR "physical* challenge*"[Title/Abstract] OR "physically challenged"[Title/Abstract] OR ("maim"[Title/Abstract] OR "maimed"[Title/Abstract] OR "maimed"[Title/Abstract] OR "maiming"[Title/Abstract]) OR ("paralyze*"[Text Word] OR "paralysis"[Text Word]) OR ("Paraplegia"[MeSH Terms] OR "Paraplegia"[Text Word] OR "paraplegic"[Text Word]) OR "quadriplegi*"[Text Word] OR ("traumatic brain injury"[Text Word] OR "brain injuries, traumatic"[MeSH Terms]) OR "acquired brain injury"[Text Word])) AND (("re-ablement"[All Fields] AND "centre"[Text Word]) OR ("reablement"[All Fields] AND "centre"[Text Word]) OR ("reablement"[All Fields] AND "hospital"[Text Word]) OR ("integrated care"[Title/Abstract:~2] OR "integrated care"[Text Word]) OR ("transitional care"[MeSH Terms] OR "transitional care"[Text Word] OR "transitional care"[Title/Abstract:~2]) OR ("intermediate care facilities"[MeSH Terms] OR "intermediate care facilities"[Text Word]) OR "reablement care"[Text Word])
58. ("amputation"[Text Word] OR "amputation, surgical"[MeSH Terms] OR ("Veterans"[MeSH Terms] OR "Veterans"[Text Word]) OR ("crippl*"[Title/Abstract] OR "disab*"[Title/Abstract] OR "handicap*"[Title/Abstract] OR ("disabled persons"[MeSH Terms] OR "disabled persons"[Text Word]) OR "physical challenge"[Title/Abstract:~2] OR "physical* challenge*"[Title/Abstract] OR "physically challenged"[Title/Abstract] OR ("maim"[Title/Abstract] OR "maimed"[Title/Abstract] OR "maimed"[Title/Abstract] OR "maiming"[Title/Abstract]) OR ("paralyze*"[Text Word] OR "paralysis"[Text Word]) OR ("Paraplegia"[MeSH Terms] OR "Paraplegia"[Text Word] OR "paraplegic"[Text Word]) OR "quadriplegi*"[Text Word] OR ("traumatic brain injury"[Text Word] OR "brain injuries, traumatic"[MeSH Terms]) OR "acquired brain injury"[Text Word])) AND (("reablement"[All Fields] AND "medicine"[Text Word]) OR (("mouth"[MeSH Terms] OR "mouth"[All Fields] OR "mouths"[All Fields] OR "mouth s"[All Fields] OR "mouthed"[All Fields] OR "mouthful"[All Fields] OR "mouthfuls"[All Fields] OR "mouthing"[All Fields]) AND "reablement"[Text Word]) OR (("occupations"[MeSH Terms] OR "occupations"[All Fields] OR "vocation"[All Fields] OR "vocations"[All Fields] OR "vocational"[All Fields]) AND "reablement"[Text Word]) OR (("cardiacs"[All Fields] OR "heart"[MeSH Terms] OR "heart"[All Fields] OR "cardiac"[All Fields]) AND "reablement"[Text Word]) OR (("neurologic"[All Fields] OR "neurological"[All Fields] OR "neurologically"[All Fields]) AND "reablement"[All Fields]) OR (("neurologic"[All Fields] OR "neurological"[All Fields] OR "neurologically"[All Fields]) AND "reablement"[All Fields]) OR (("psychiatrical"[All Fields] OR "psychiatrically"[All Fields] OR "psychiatrics"[All Fields] OR "psychiatry"[MeSH Terms] OR "psychiatry"[All Fields] OR "psychiatric"[All Fields]) AND "reablement"[Text Word]) OR ("reablement"[All Fields] AND "nursing"[Text Word]) OR (("stroke"[MeSH Terms] OR "stroke"[All Fields] OR "strokes"[All Fields] OR "stroke s"[All Fields]) AND "reablement"[Text Word]))
59. ("amputation"[Text Word] OR "amputation, surgical"[MeSH Terms] OR ("Veterans"[MeSH Terms] OR "Veterans"[Text Word]) OR ("crippl*"[Title/Abstract] OR

"disab*"[Title/Abstract] OR "handicap*"[Title/Abstract] OR ("disabled persons"[MeSH Terms] OR "disabled persons"[Text Word]) OR "physical challenge"[Title/Abstract:~2] OR "physical* challenge*"[Title/Abstract] OR "physically challenged"[Title/Abstract] OR ("maim"[Title/Abstract] OR "maimed"[Title/Abstract] OR "maimed"[Title/Abstract] OR "maiming"[Title/Abstract]) OR ("paralyze*"[Text Word] OR "paralysis"[Text Word]) OR ("Paraplegia"[MeSH Terms] OR "Paraplegia"[Text Word] OR "paraplegic"[Text Word]) OR "quadriplegi*"[Text Word] OR ("traumatic brain injury"[Text Word] OR "brain injuries, traumatic"[MeSH Terms]) OR "acquired brain injury"[Text Word])) AND "reablement"[All Fields]

60. "reablement"[Text Word]
